# How organizations can support interprofessional collaboration through the eyes of faculty and other professionals

**DOI:** 10.64898/2025.12.02.25341493

**Authors:** Maura N Polansky, Arielle L. Buslovich, Joyce Maring, Anne Thompson

## Abstract

**Introduction:** Interprofessional collaborative practice (IPC) is deemed essential to address the quadruple aims of improving healthcare. There is also emerging interest in the role IPC among healthcare faculty in their educational and research roles educators. Various facilitators and barriers of IPC at the individual and team levels have been identified. However, organizations and systems are often strong drivers for professional practice and limited attention has been placed on the strategies needed for organizations to support IPC. This study explored the perspectives of faculty and other professions regarding organizational facilitators and barriers to IPC, as well as strategies to support IPC within their own professional contexts.

**Methods:** Document analysis was performed on papers from a course entitled “Interprofessional Collaboration in Practice”, a requirement of a Doctor of Health Sciences in Clinical and Academic Leadership program. The papers were written as part of a culminating project exploring opportunities to enhance IPC within students’ own professional contexts. Students used reflections of their own professional experiences, informed by the literature and other course content, as well as informal interviews with other professionals in their workplace, to explore facilitators, and barriers to IPC. Inductive content analysis was performed by two members of the research team. Final results and selective quotes were reviewed by the full research team.

**Results:** Twenty papers written by students of various professional backgrounds, professional roles, and organizational types were included.

Four dominant themes were identified that reflected facilitators and barriers to IPC that existed within students’ organizations. These four aspects of organizations appear to be both essential and inter-related, and include organizational systems, strategic priorities, culture, and institutional leaders.

**Conclusion:** Organizations and their senior leaders play a critical role in developing collaborative organizations where employees from various professions effectively work together to accomplish organizational goals. In addition to the current practice of preparing individual healthcare professionals for IPC, attention to organizations and their leaders is essential. Faculty and other professionals have particularly valuable perspectives in understanding how IPC can be advanced in healthcare organizations and should be the focus of future professional development initiatives and additional research related to IPC.

**Trial registration:** not applicable

## Introduction

Interprofessional collaborative practice (IPC) is deemed essential for addressing the quadruple aims in healthcare delivery.^1,2^ There is also emerging interest in the role IPC among healthcare faculty in their educational and research roles.^3,4^ Although IPC has shown significant benefits to organizational efficiency, patient care, and patient safety, IPC has not been extensively adopted throughout all clinical settings and little is known about its role in non-clinical health-related organizations.^5^

The adoption of collaborative practice in healthcare, from more traditional approaches, requires a major paradigm shift.^6^ The lack of IPC adoption has been perceived as being primarily rooted in the siloed approach to health professions education (HPE), where students from each profession are taught separately.^1^ This pedagogical approach to HPE is thought to result in a lack of development of essential competencies for IPC and may even lead to the development of biases and negative stereotypes towards those from other professions.^7^ Therefore, the primary approach adopted to support IPC in healthcare has been through changes in HPE. ^8^

Most attention to changing HPE in support of IPC has been focused on the education of new healthcare professionals. Extensive efforts have been devoted to the preparation of a collaborative practice-ready workforce through the integration of interprofessional education (IPE) in HPE programs.^9^ IPE is defined as individuals from two or more professions learning with, from, and about each other.^9^ IPE aims to develop competencies for interprofessional practice while also supporting positive attitudes and beliefs about other professions and the value of collaborative practice.^8^ This novel pedagogy is now commonly integrated into profession-specific healthcare programs such as pre-licensure schools and post-licensure residencies. In fact, for the last few decades, IPE has been increasingly required by U.S. specialty accrediting bodies for health professions programs, including those in medicine, nursing, physical therapy, and pharmacy.^10,11^

Despite the attention placed on IPC competency development for healthcare professionals and the routine use of IPE to develop these competencies, the full adoption of effective IPC has not been fully realized. Impediments to IPC may be challenging to overcome, especially by professionals new to the workplace, and despite the development of their interprofessional competencies through prior IPE experiences. While numerous scholars have explored barriers to collaboration that occur at the individual and team levels, limited attention has been placed on how barriers at the organizational level may impede IPC and effective organizational strategies to overcome these barriers.^12^

Organizations and systems are often strong drivers for professional practice.^13^ In healthcare-related organizations, attention to patient safety and healthcare quality, such as through the development of high-reliability organizations, has been recently emphasized.^14^ However, limited attention is given to how organizations can facilitate or impede IPC to support high reliability. A recent systematic meta-review by Wei and colleagues (2022)^12^ identified organizational factors, including values, culture, structure, and support, that may influence IPC. Despite these findings regarding organizational influences on IPC, strategies to overcome organizational barriers have been largely overlooked.

With the primary focus on the training of new healthcare professionals, limited attention has been placed on the role of faculty development and other professional development programs that support organizational change. Emerging leaders in healthcare-related workplaces, including but not limited to health science faculty, may be uniquely positioned to recognize organizational factors that impact IPC. These professionals typically engage with others at various levels of the organizational hierarchy and across multiple units. Furthermore, emerging leaders, particularly those pursuing professional development opportunities in healthcare leadership, are likely primed through their educational experiences to identify strategies to overcome organizational barriers to IPC and may be uniquely positioned to impact organizational change.

Graduate programs that exist to develop expanded expertise for healthcare professionals (typically providing second graduate degrees) enroll faculty and other professionals from various health professions who hold early- and mid-career leadership positions or aspire to leadership positions in clinical, academic, and other healthcare-related organizations.^15,16^ As such, “the mission of most of these programs is to prepare leaders in the health professions who can manage change within their institutions, overcome organizational barriers, and effectively direct the future of health care delivery systems”.^15^ Therefore, we sought to explore the perspectives of faculty and other emerging leaders in healthcare-related organizations pursuing graduate studies in order to identify organizational facilitators and barriers to IPC, as well as potential strategies to overcome these barriers.

## Methods

### Context

For this study, a Doctor of Health Sciences in Clinical and Academic Leadership program was chosen. Students enrolled in this program come from a wide variety of professional backgrounds, including clinical professions (e.g., nursing, medicine, dentistry, physical therapy), laboratory and basics sciences, and other related fields, including public health and health care administration. This part-time program typically enrolls students who are employed full-time or part-time while enrolled in the program. This work experience provides them with an opportunity to apply course content to their own place of authentic professional work for program coursework and assignments. Students are employed in various educational and healthcare institutions across the United States, including faculty positions and those in other professional roles. As emerging leaders, students are or will be responsible for the educational and professional development of others and may serve as leaders of clinical and non-clinical activities related to effective IPC. As early to mid-career professions, they were expected to be uniquely positioned to observe and examine the organizational characteristics at play in their own professional contexts.

The selected graduate program was identified in part due to its requirement that all students complete a course on IPC entitled “Interprofessional Collaboration in Practice”. The course was developed using a design-based research approach as described by Dolmans and Tigelaar (2012).^17^ This approach to course development requires the utilization of theory to guide curriculum development and evaluation, providing opportunities for research to be conducted during the design or implementation phases to advance relevant theory. The development of this course using the design-based research approach has been previously described.^18^ (Polansky 2022)

As a major course project and final paper, students were asked to explore opportunities to enhance IPC within their own professional context and to identify facilitators and barriers to IPC that currently exist. Students were also expected to consider potential strategies to overcome those barriers. They used the course content (readings, assignments, and class discussions) to guide this work and were required to discuss their project with other professionals within their organization through informal interviews and/or focus groups. These course papers provided an opportunity for the research team to analyze students’ perspectives.

### Study design

A qualitative document analysis of the final course papers was performed. Course papers were drawn from students who completed the course across two semesters in 2021 and 2022 at the George Washington University. A total of 20 papers were available for analysis.

### Data Collection and Management

Background information about each student was collected as part of the course introductions. The “Interprofessional Collaboration in Practice” course was taught by the study PI (MNP), who de-identified each paper for the research team. Therefore, the identity of the student associated with their respective paper was only known to the PI.

Study data was stored in a password-protected online content management system (i.e., Box). A qualitative analysis software (VERBI Software MAXQDA 24) was used for data management.

### Data Analysis

Document analysis was performed on each of the 20 student course papers. Document analysis is the process of analyzing documents to “uncover meaning, gain understanding, and come to conclusions”.^19^ The analysis was guided by processes of first and second cycle coding as described by Saldaña (2016). ^20^

Inductive document analysis was conducted by two members of the research team (MNP, ALB) through multiple readings of the course papers, followed by open coding and the development of key themes.^21^ (Elo & Kyngas, 2008). First-cycle coding was performed with line-by-line coding to provide close analysis of each document. Initial codes were generated that described facilitators and barriers to IPC within the writers’ professional contexts. The researchers met regularly to discuss the analysis, and the grouping of codes was performed together. Next, second-cycle coding was performed to identify key themes related to organizational facilitators and barriers. This analysis continued until a sufficiency of themes were identified (i.e., information power).^22^ The full research team reviewed selective quotes and the final thematic results.

Analytic memo writing was performed throughout the process to support the development of key themes as they were identified and to establish higher-order codes.^23^

## Positionality and reflectivity

All members of the research team have experience in interprofessional clinical practice as current or former healthcare professionals (physician assistant, occupational therapist and physical therapists).

The primary investigator (MNP) served as the course director, both developing and providing instruction for the course during the semesters from which the course papers were drawn from. As such, MNP was aware of the students who wrote each paper through their enrollment in the course. MNP has experience in IPC and IPE in various clinical and academic settings and has expertise in the emerging literature on IPC.

Co-investigator ALB was a graduate student in a clinical health professions program, during the data analysis phase of the study. She was also involved in data analysis, as discussed below.

Co-investigator JM has extensive experience in interprofessional clinical and academic collaborative practice. She was the founding program director of the respective program at the time the students were enrolled in the course. However, JM was not involved in reviewing course papers and had only access to selective quotes.

Co-investigator AT served as the senior author, given her extensive expertise in IPC. She also provides instruction in topics of interprofessional practice to graduate students in HPE at another university.

Reflectivity was performed through regular discussions during research meetings to enhance the trustworthiness of the findings. Reflexivity has been defined as “the process of examining both oneself as researcher, and the research relationship” and consists of exploring one’s assumptions and preconceptions and considering how these may affect research decisions.^24^ Reflexivity was conducted through discussions among the research team about their viewpoints and to challenge potential biases as they emerged throughout the study. Reflexivity also was conducted by reflective memo writing by the primary investigator (MNP).

## Data Availability

The de-identified data that support the findings of this study are available from the primary author, but restrictions apply to the availability of these data, and they are not publicly available. The data are available upon request and with the permission of George Washington University review board.

## Declarations

### Ethics approval and consent to participate

The research protocol was deemed exempt by the George Washington University institutional review board in October 2021. The study adhered to the Declaration of Helsinki ethical principles.

Formal informed consent was deemed unnecessary by the institutional review board. Instead, students enrolling in the course were notified that their de-identified data may be used for research purposes and were provided the opportunity to decline use of their data. No students declined data utilization.

Due to the nature of limited student enrollment in the specific program, all quotes will be referred to by student number only to avoid reveling identifying information.

### Consent for publication

Not applicable.

## Funding

No external funding was provided for this research.

### Authors’ contributions

MNP - made substantial contributions to the conception, design, acquisition, analysis, interpretation of data and drafted the work.

ALB- made substantial contributions to the analysis and interpretation of data and substantively revised the manuscript.

JM- made contributions to the conception of the work and substantively revised the manuscript.

AT- made contributions to the conception of the work and substantively revised the manuscript.

All authors approved the submitted version and agree to both be personally accountable for their own contributions and to ensure that questions related to the accuracy or integrity of any part of the work, even ones in which the author was not personally involved, are appropriately investigated, resolved, and the resolution documented in the literature.

## Acknowledgements

The research team would like to acknowledge the initial design-based research team who contributed to the development of the respective course. This includes Debra Herrmann, Anthony R. Artino Jr, and Ryan Strauss at the George Washington University.

We also expression our sincere appreciation to students enrolled in the course for their willingness to allow the use of their data for research purposes.

## Results

### Background of students

Students whose written course papers were included in the study included faculty from health sciences programs, as well as other professionals at various health-related organizations. Students’ background professions included various clinical professions, public health, basic sciences, and administration. Many held faculty or adjunct faculty positions while others served in various roles such as managers, coordinators, team leads, and research specialists or liaisons. Senior leaders were also represented.

Students were employed in various educational settings including at universities, community college and pre-health professions programs. Other students were employed by hospitals, governmental agencies and for-profit companies. Only limited student data in aggregate is reported here to ensure anonymity.

### Potential impact of enhanced IPC

Students discussed a wide variety of opportunities for IPC within their organizations, as well as the potential positive impact of more effective IPC practices. Enhancing educational programs involving health professions students and clinical trainees was discussed in many papers (P1, 2, 3, 5, 6, 12, 13, 14). For students involved in clinical practice, enhanced IPC was identified as a means of improving patient outcomes (P 1, 5, 9-11, 13, 14, 16, 18-20). Increased opportunities for research, publication, and grantsmanship were also discussed by faculty and other professionals involved in research (P1, 4, 5, 6, 8, 15, 17). Improving efficiencies, such as sharing or securing necessary resources, collaborative teaching, enhancing case management, and developing clinical policies were also identified (P 1, 2, 10, 13, 14, 20). Some papers included reflections on lasting organizational change that was expected to result from enhanced IPC (P 1, 14, 16, 17).

### Organizational facilitators and barriers

Four major themes related to organizational facilitators and/or barriers to IPC were identified in the papers analyzed. They include organizational systems, strategic priorities, organizational culture, and institutional leaders. These four organizational domains appear to be both essential and inter-related. (See Figure 1.)

> “These strategies (to support IPC) are interdependent. Some strategies can be barriers, if not applied prudently and judiciously” (P6)

> “No single strategy can lead to the adoption of IPC, but by using multiple strategies to mitigate the identified barriers, IPC may be adopted as standard practice…” (P10)

**Figure 1.**
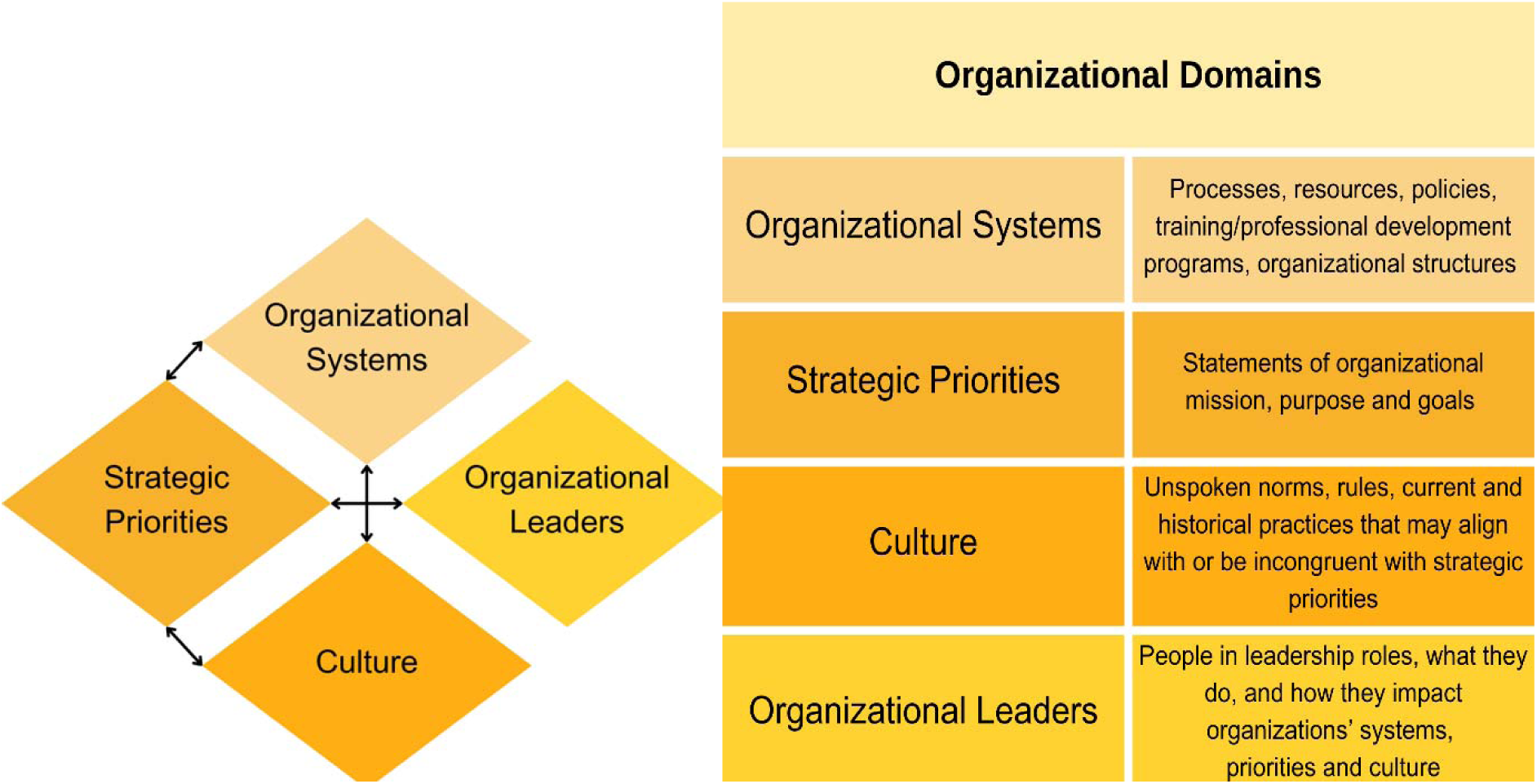
Four essential and interrelated organizational domains influencing IPC

**Organizational systems, including processes, resources, policies, employee development programs, and other organizational structures,** were extensively discussed across the students’ papers as facilitators or barriers to IPC.

Numerous papers addressed the challenges that often exist when professionals from different professions are also from different organizational units, such as different offices or departments, clinical wards, universities, colleges, and service committees. Having structures to bridge these silos and having formal opportunities to gather across units were seen as essential strategies. Having dedicated time and physical space for IPC was noted to be particularly important for IPC to occur, as well as essential in providing opportunities for professionals to build foundational relationships.

> “The nurse’s station is the central place where interprofessional interactions occur. For instance, when needing to interact with other doctors or nurses, the nurse’s station tends to be the meeting point. When the lab calls the ward, the contact tends to go to the nurse’s station before reaching the patients’ nurse.” (P11)

The lack of co-location by many professionals involved in common work was a frequently identified barrier for IPC. This included those working in different professional locations and with remote work arrangements.

> “In an ideal world, the entire team would work in the same location, sharing information about each patient and working collaboratively on the best care plan.” (P20)

Working in different settings was perceived as inferior to in-person collaboration but could be partially overcome with the help of dedicated systems, such as the use of technology for virtual meetings.

> “Since we are a hybrid team, it is important to have communication tools that are available to everyone, in real-time to assist with some of those communication gaps that occur within a dispersed team.” (P20)

Various types of meetings were mentioned for fostering IPC. Formal meetings supported learning about other professionals’ roles and responsibilities, discussing goals, and coming to shared understandings. Meetings were perceived as providing opportunities for professionals to understand the commonalities and differences that exist between professions, including those related to professional competencies and roles. Moreover, opportunities for informal meetings could support relationship building, open communication, and getting to know each other personally, which in turn could further improve IPC.

While IPC was typically seen as occurring organically, some papers discussed the importance of planning for and even managing IPC by unit leaders. This required dedicating time and establishing formal systems for IPC planning.

> “By establishing collaboration as a step on the checklist, this tool can be used to help emphasize the need and benefit of IPC.” (P10)

Most papers indicated that there was limited or no attention to developing the competencies for effective IPC within their organizations.

> “Within our organization, there currently are no faculty development programs focused on teamwork and collaboration.” (P3)

Organizations could support IPC by providing training and education for their workforce to develop professionals’ skills in IPC. Providing professional development opportunities in the workplace, such as incorporating topics related to IPC into new employee/faculty orientations or required training programs, was discussed as a potential strategy to support the development of competencies for IPC.

> “A methodology to build or rebuild an interprofessional team must begin with education and training.” (P19)

> “Implementation of professional development on IPC and IPE theory and best practices will help faculty to become more aware of the silos that exist within our programs, so that we can actively work to break them down.” (P3)

Some papers discussed the lack of established incentives for IPC and how developing incentives for effective IPC could be of value within the organization.

> “Participants [interviewed by the student during the course] spoke to the institution lacking strong incentives to collaborate.” (P5)

Finally, some papers discussed the importance of having institutional policies that directly address IPC. For example, developing institutional policies that require broad professional representation for organizational activities and responsibilities was perceived as valuable.

> “All these professions are represented at the organization’s [policy] committee, which approves all organizational policies.” (P10)

## Strategic priorities

Several papers discussed the importance of strategic priorities related to IPC for organizations (or major units within the larger organization). Strategic priorities included **explicit statements of organizational mission, purpose, and goals that aligned directly or indirectly with IPC.**

Some papers indicated that organizations had explicit missions, visions, and/or strategic priorities addressing IPC.

> “Collaboration is encouraged as outlined at all levels within the university though mission statements and departmental goals and objectives. This creates a positive environment to collaborate among programs, departments, and school.” (P5)

Furthermore, when IPC is prioritized at the organizational level, more time, effort, and resources were dedicated to improving collaboration.

> “They [professionals interviewed by the student] spoke to inclusion of interprofessional collaboration in the strategic plan as a mechanism to provide a shared goal for the school and a way to create incentives for IPC.” (P15)

Other papers discussed how IPC was more implicit within their organization’s strategic agenda. For example, IPC was often perceived as a means of accomplishing institutional goals, such as providing quality education or healthcare.

> “The mission speaks to the pursuit of excellence in educating a diverse workforce and leaders in health professions”. (P5)

> “Our overarching goal is to have every member of our team work together to prevent chronic diseases and encourage healthy behavior change throughout our organization” (P20)

In contrast, the lack of strategic priorities related to IPC was noted by authors of other papers and seen as a major barrier to IPC. As such, addressing IPC within strategic planning was perceived as an important step in supporting organizational change towards enhanced IPC.

> “A commitment must be made to reach the goal of interprofessional collaboration at our agency.” (P16)

> “Faculty in the focus group spoke to institutional change as a method to improve interprofessional collaboration. They spoke to inclusion of interprofessional collaboration in the strategic plan as a mechanism to provide a shared goal for the school and a way to create incentives for IPC.” (P5)

## Culture

The overall culture of the organization was also viewed as a crucial domain, either as a facilitator or an impediment to IPC, and papers discussed how cultural change may be necessary to enhance IPC. Organizational culture was reflected in comments about **unspoken norms, rules, and practices, including historical practices, that may align with or be incongruent with organizational priorities.**

Some papers discussed how their organization had a strong commitment to or a long-standing history of IPC, which was apparent when they joined the organization. This culture of IPC fostered further collaborations.

> “According to the interviewed … faculty, there was already a history of joint research activities between two professions… Prior positive collaborative experiences between [two specific professions] demonstrate that the spirit of teamwork between the two programs already exists. Sharing this history with faculty in either profession who are unaware of it may stimulate greater enthusiasm and spur new ideas for collaboration.” (P6)

Yet other students discussed the obvious lack of an organizational culture to support IPC.

> “The organization’s culture has not placed an emphasis on the importance or benefits of IPC” (P10)

Furthermore, many students discussed how a culture of professional hierarchies at various levels of the organization was apparent; and despite any explicit or implicit messages regarding professional inclusion, the acceptance of working in silos was noted to exist.

> “The nurses (interviewed by the student) expressed their level of dissatisfaction with the attitude and culture of the organization that condones and accepts a hierarchal structure.” (P19)

Papers shared reflections on the need for cultural change, facilitated by organizational leaders to overcome barriers IPC.

## Organizational Leaders

Course papers reflected on the important role of organizational leaders in creating the conditions or barriers for IPC. Leaders, **the people in leadership roles, what they do, and how they impact the organizations’ systems, priorities, and cultures** were discussed in many papers. These leaders included students’ immediate supervisors, as well as those at the organizational level, such as senior administrators. Some students also reflected on their own current and future roles as leaders and how they could potentially impact conditions that facilitate IPC in their organization.

First, leaders were perceived as needing to demonstrate the value of IPC through their own actions. Selecting leaders with a commitment to and skills in IPC was identified as an important strategy to support organizational change.

> “These actions can only occur if leadership from the top-down buys into these principles and promotes this culture through workshops, trainings, and meetings with staff.” (P16)

Having established support from leaders for IPC initiatives was also seen as imperative.

> “Strong leadership is also an important organizational facilitator of collaboration. All interviewed faculty pointed out that they do already have the support of higher leadership -deans and department chairs -to embark on joint interprofessional ventures.” (P6)

Support from leaders included providing others with dedicated time for collaborative work, as well as providing incentives to encourage collaboration.

> “It’s also a potential resolution to discuss with the PDs (program directors) whether it would be possible to have the faculty members have reduced workloads which would help them collaborate more efficiently…” (P1)

Leaders’ actions also included demonstrating effective IPC through their own actions, such as establishing trust and transparency, the inclusion of all professions and ensuring each profession has an equal voice, and managing interprofessional conflict, including facilitating the resolution of long-standing interprofessional problems.

> “Constructively addressing conflict, a culture of transparency, holding each team member accountable for their actions, and objective and transparent productivity metrics will lead to a cohesive and effective [interprofessional] team.” (P15)

It was observed that leaders themselves should be collaborative and foster collaboration within their own leadership teams, as well as across the organization.

> “Leadership needs to be collaborative in nature with shared responsibility.” (P3)

> “An important first step for all parties is for leadership to take initiative in collaborating so that the workplace can learn about each other’s professions.” (P16)

Some students also discussed the value of leadership providing support for IPC initiatives led by others.

> “The medical program PD explained how to address the committee and that he would like to review my proposal to the committee so that it has the highest chance of success of approval.” (P1)

### Students as leaders (Owning it)

Some participants reflected on their own roles as leaders within their organizations and their roles as current or future leaders in facilitating IPC. Through their own leadership actions, students recognized the need to build trust across professions and foster relationships with other organizational leaders from various professions, demonstrating the value of IPC in achieving organizational goals.

> “My current role as an [faculty member and director] lends itself well to multiple opportunities for IPC among faculty.” (P5)

Some reflected on the importance of their actions as a leader in overcoming barriers to IPC and potential strategies to do so.

> “My job is to develop strategies that to deal with and prevent conflict and keep our team productive and patient focused.” (P20)

> “Quarterly, I plan to distribute a feedback survey that discusses any challenges associated with unpredictability, unproductiveness, limited collaboration, and conflict. Hopefully, this will assist with conflict but also break down some of those unapproved hierarchical roles that breed conflict.” (P20)

The need for further their own professional development was also noted as a mean to enhance their skills related to IPC, such as in managing interprofessional conflicts.

> “Ultimately, being in a position of power and influence, helps to shift the culture focus to one that is about teamwork and collaboration…After identifying gaps within my own leadership abilities, I can work towards developing skills in those areas” “By being aware of, and willing to grow my own leadership abilities, I will be able to know when and what method to apply to make sure our leadership remains collaborative.” (P3)

## Discussion

This study of faculty and other emerging leaders in the health sciences highlights how various domains of organizations can either facilitate or hinder interprofessional practice. The organizational domains identified include organizational systems, strategic planning, culture, and organizational leaders; these domains are both essential and inter-related providing a framework to examine organizational influences on IPC. Of note, this framework aligns well with other organizational frameworks in the literature, including organizational factors (values, culture, structure and support) found to be barriers to IPC in the work by Wei et al (2022)^12^ and the widely used Bolman and Deal (2013)^25^ leadership framework (human resource, structural, political and symbolic). However, our study also illuminates specific strategies in healthcare-related organizations that may be used to address systems barriers to IPC.

The organizational strategies to address barriers to IPC included numerous potential approaches. Examples include having IPC explicitly addressed in statements of mission, values, and priorities; creating formal opportunities for meeting across organizational units; providing time, physical spaces, resources, and financial incentives for IPC; providing professional development initiatives within the workplace regarding IPC; and ensuring institutional leaders demonstrate the value of IPC through their own actions. Of interest was the priority students placed on the need for their own leaders-supervisors, and those in high-level positions -within the organization to support IPC initiatives proposed by them and their colleague. Many students also reflected in their writings on their own role as current or future leaders in supporting the advancement of IPC and the importance of further professional development to support their skills in IPC. These results provide further weight to the notion that the use of IPE with early learners is likely insufficient by itself in creating the paradigm shift necessary to develop effective collaborative professional environments.

The utilization of the graduate student course papers for context analysis, as a research method supported by the use of design-based research, provided rich data to explore the research topic. As anticipated, course content and learning activities, and the experiences of other professionals whom the students selected to interview for their projects, primed students to recognize organizational influences in their professional context. Furthermore, students’ positions as emerging leaders allowed them to see professions at all levels of their organization, including senior-level leaders as well as those who do not hold formal leadership positions, through their daily work.

With the results of this study highlight the importance of organizational support for IPC, the role of leaders at all organizational levels should not be understated. Leaders are increasingly recognized as critical to the advancement of interprofessional practice in both clinical and academic settings^.18^ However, emerging leaders and mid-career HPE faculty are not frequently the focus of professional development activities including those aimed at advancing in IPC.^26^ This is despite their unique roles and perspectives which provide important opportunities to advance IPC efforts in both research and practice.

Faculty and other professional development programs, including initiatives in healthcare leadership and health professions education, may provide unique opportunities to support IPC.^16^This study also illustrated the potential role of graduate programs, one form of faculty and professional development, in advancing IPC. Such programs will likely be more effective if they are themselves interprofessional learning environments that foster professional inclusivity.^16^ The inclusion of curricula in such programs that focus on interprofessional practice and learning may provide valuable opportunities to support faculty/professional development for emerging leaders to advance the necessary paradigm change that has so far been elusive.

### Limitations

While the results of this study bring important insights to the body of literature regarding IPC, some limitations exist. As with all qualitative research, this study aimed to deeply explore the perspectives of a small number of individuals with diverse professional experiences. As such, the perspectives of the writers represented in this study may be different from other professionals in healthcare related organizations. In particular, the perspectives of senior leaders and employees with limited leadership experience or responsibilities in the workplace may vary considerably from those represented in this study. However, these findings may be transferrable to similar professionals, such as other faculty and mid-career professionals.

This study used the method of content analysis to examine papers written as part of a required course assignment. As such, students’ writings may have been influenced by their perceived expectations of their instructors. For example, students may have over or under emphasized or omitted the role of some conditions related to IPC.

Despite these limitations, these study findings should be instrumental for future efforts to foster effective IPC practices.

### Future directions

Based on the results of this study as well as previously published research on IPC, the authors provide the following recommendations.

First, health-care related organizations should attend to the critical role of organizational culture, systems, and strategic planning in advancing efforts towards IPC. Leaders at all organizational levels should develop their own competencies in IPC, ensure they model effective collaboration within their own teams and across organizational units, and seek opportunities to support the efforts of others in advancing IPC.

Second, interprofessional faculty development and other professional development opportunities should be provided to professionals in leadership and non-leadership roles to support the development of a collaborative workforce. They should also address change management at the organizational level related to Interprofessionalism.

Finally, further research on IPC should focus on faculty and professional development initiatives, leadership, and organizational change as critical topics to support the necessary paradigm change for the broad adoption of effective IPC.

## Conclusions

Organizations and their leaders play a crucial role in fostering collaborative environments where employees from diverse professions effectively work together to achieve organizational goals. In addition to the current practice of preparing individual healthcare professionals for IPC, attention to organizations and their leaders is also essential. Emerging leaders and mid-level professionals, such as faculty and managers, have particularly valuable perspectives in understanding how IPC can be advanced in healthcare organizations and should be the focus of future professional development initiatives and additional research related to IPC.

